# Guided versus standard antiplatelet therapy in patients undergoing interventional intracranial aneurysm treatment using stents: protocol of a cluster randomized controlled cohort study

**DOI:** 10.1101/2023.11.16.23298664

**Authors:** Yangyang Zhou, Wenqiang Li, Hongqi Zhang, Bin Luo, Yang Wang, Jun Wang, Pinyuan Zhang, Liang Li, Fushun Xiao, Shiqing Mu, Jian Liu, Yuanli Zhao, Xinjian Yang, Peng Jiang

## Abstract

**Background:** The goal of standard dual antiplatelet therapy (SDAT) in patients undergoing endovascular interventional treatment of intracranial aneurysms is to prevent thrombosis; however, some patients have a poor response to these drugs, which increases the risk of cerebral infarction. This study aims to examine whether adjusting antiplatelet therapy based on light transmission aggregometry (LTA) can reduce the incidence of ischemic events compared with SDAT.

**Methods:** We will conduct a cluster randomized controlled trial using 16 treatment teams from eight hospitals in mainland China, enrolling 590 patients with unruptured intracranial aneurysms treated using endovascular stent placement. The treatment teams serving as clusters will be randomly assigned to either the test or control group at a 1:1 ratio. Test group patients will receive an antiplatelet regimen guided by LTA. Control group patients will receive SDAT. Patients will be followed for 1 month after the treatment period. The primary outcome measure is cerebral ischemic events within 30 days of stent placement, including stent thrombosis, ischemic stroke, and transient ischemic attack. The safety measure is all bleeding events within 30 days of treatment.

**Discussion:** The trial aims to determine whether LTA-guided antiplatelet therapy reduces the incidence of ischemic events without increasing the risk of bleeding in patients with intracranial aneurysms treated with endovascular stenting. Completion of this clinical trial may provide an individualized safe and effective regimen for antiplatelet therapy.

Trial registration ClinicalTrials.gov, NCT05825391. Registered on April 11, 2023.

**What is already known on this topic:** Dual antiplatelet therapy is administered to reduce thrombotic events in neurointerventional therapy of intracranial aneurysms. Platelet function testing is used to evaluate the antiplatelet effect of aspirin and clopidogrel.

**What this study adds:** The trial aims to determine whether LTA-guided antiplatelet therapy reduces the incidence of ischemic events without increasing the risk of bleeding in patients with intracranial aneurysms treated with endovascular stenting.

**How this study might affect research, practice or policy:** For stent neurointerventional therapy of unruptured intracranial aneurysms, this study could provide a promising method for adjusting appropriate antiplatelet therapy.

## Background

Intracranial aneurysms are generally treated using endovascular interventional therapy when possible.^1,2^ The use of intracranial endovascular stents, particularly flow diverters, has greatly expanded the indications for interventional therapy and has contributed to improved treatment success rates. However, the presence of an endovascular stent also increases the risk of ischemic complications.^3,4^ Standard dual antiplatelet therapy (SDAT), which consists of oral aspirin 100 mg and clopidogrel 75 mg daily, is administered to stent patients to reduce the incidence of thrombotic events. However, the response to antiplatelet agents varies between patients. Some patients respond poorly, particularly to clopidogrel, and the incidence of thrombotic events can reach 40%.^5^

Platelet function testing is important to identify patients with antiplatelet drug resistance. Intensifying antiplatelet therapy in such patients may help reduce the risk of ischemic complications.^6,7^ Light transmittance aggregometry (LTA) is the gold standard for detecting platelet aggregation function, which may have importance in patients undergoing interventional intracranial aneurysm treatment.^8^ Maximum platelet aggregation rate induced by adenosine diphosphate (ADP) can be used to predict ischemic events using previously established cut-off values.^9^

Ticagrelor is a reversible P2Y12 receptor antagonist with rapid onset. Unlike clopidogrel, it does not require CYP450 enzymatic metabolism for activity and its efficacy is not affected by CYP2C19 gene polymorphisms.^10^ Therefore, we will conduct a cluster randomized controlled cohort study to examine whether LTA-guided antiplatelet therapy reduces the incidence of ischemic complications in patients undergoing endovascular intervention for intracranial aneurysm treatment.

## Methods

### Study design and objectives

This is a prospective multicenter cluster randomized controlled study evaluating the efficacy and safety of LTA-guided antiplatelet therapy in patients with unruptured intracranial aneurysms undergoing endovascular stenting. Detailed trial protocol and statistical analysis plan are available in **Supplement 1**. Sixteen neurointerventional treatment teams from eight hospitals will be involved. Treatment team inclusion criteria are shown in **Box 1**. The 16 teams will be randomly divided into test and control groups at a 1:1 ratio. Test group patients will receive an antiplatelet regimen guided by LTA testing and control group patients will receive SDAT. The primary outcome measure will be incidence of ischemic events 30 days after treatment, which will be compared between the test and control groups (**Fig. 1**).

**Figure 1.**
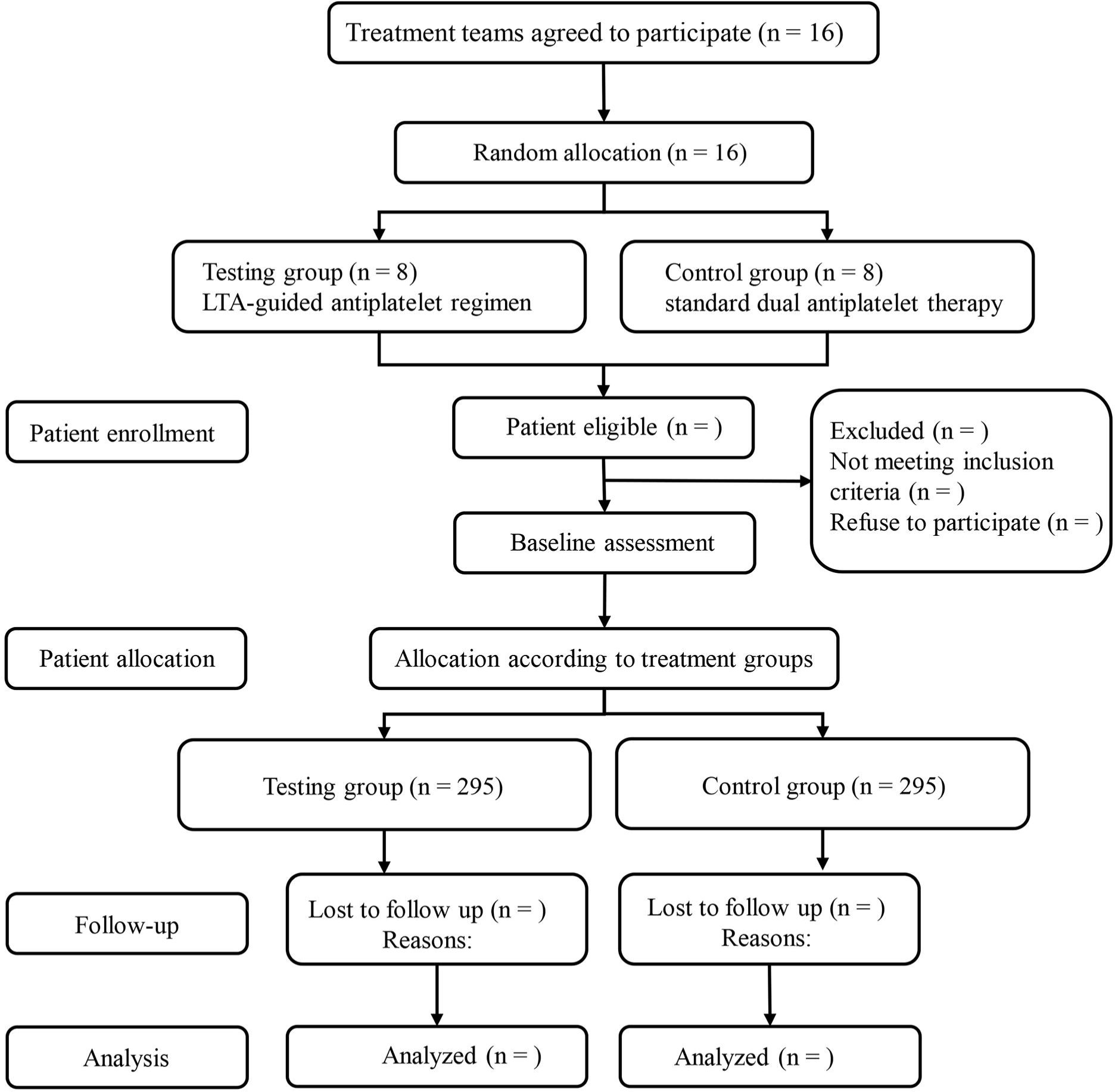
Study flow chart. LTA, light transmittance aggregometry.

**Box 1.**
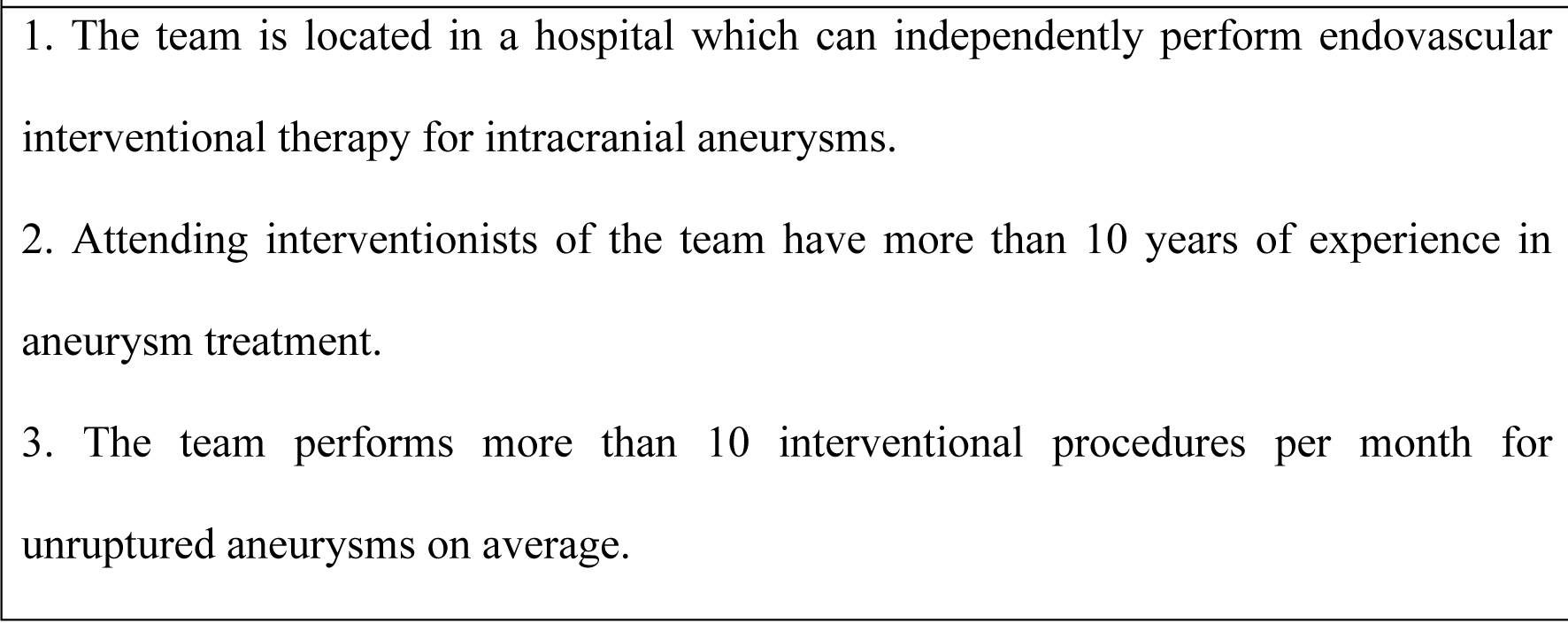
Neurointerventional treatment team inclusion criteria.

### Study population

Patient inclusion and exclusion criteria are shown in **Box 2**. The study will enroll 590 patients with unruptured intracranial aneurysms.

**Box 2.**
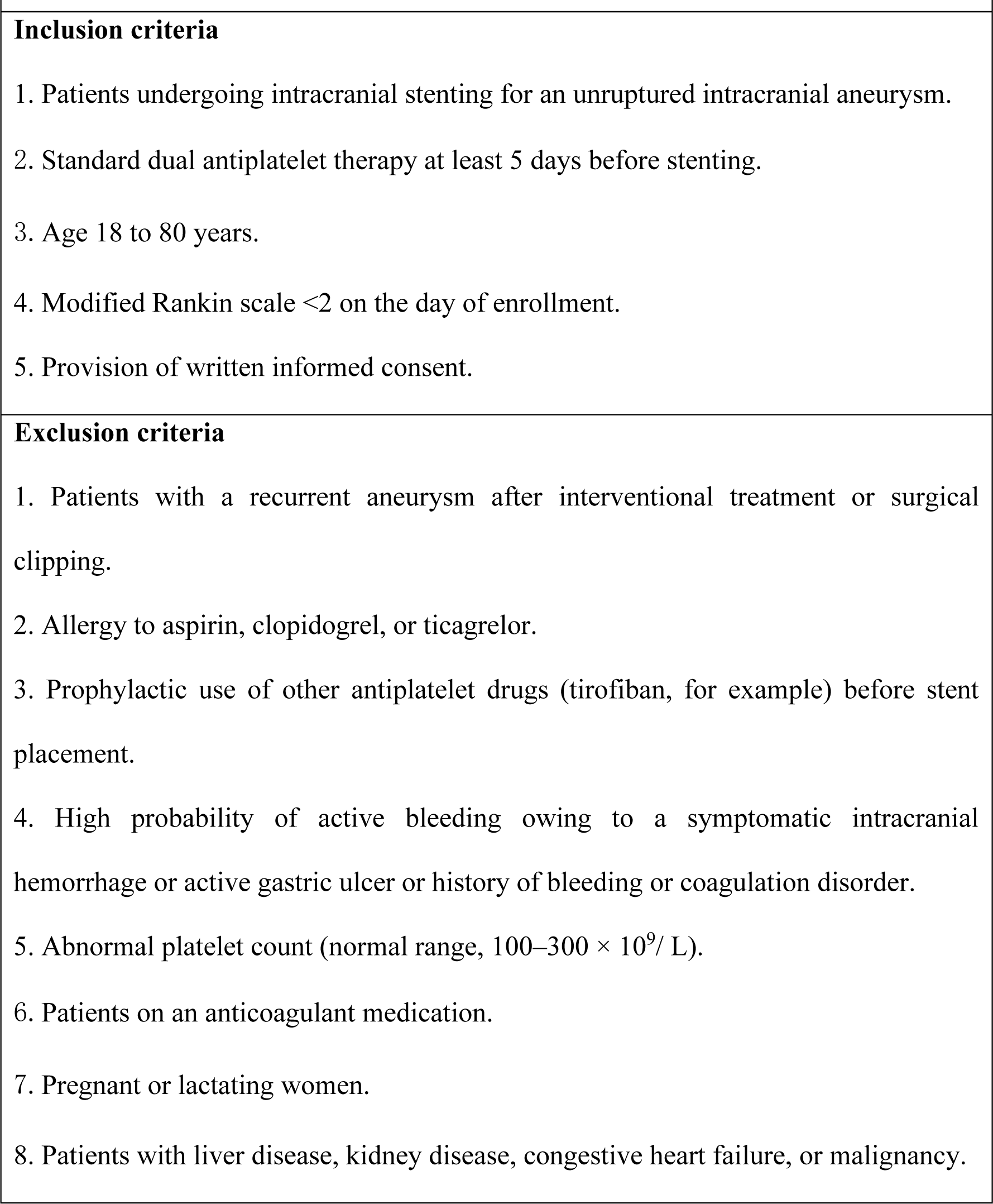
Patient inclusion and exclusion criteria.

### Measures and observation variables

We will collect the following data: 1) demographic characteristics—age, gender, height, weight, body mass index; 2) past medical history—hypertension, diabetes, coronary heart disease, hyperlipidemia, medications (type, dose, frequency), smoking and drinking history, family history, surgical history; 3) laboratory testing data— complete blood count, liver function, kidney function, blood glucose, blood lipids, coagulation parameters, LTA; 4) physical examination—neurological examination, modified Rankin scale score; 5) aneurysm characteristics—size, number, location, shape, neck width, diameter of parent artery.

### Randomization

Random numbers will be generated by an independent third-party using SPSS software version 26.0 (IBM Corp., Armonk, NY, USA) to generate a random sequence. Eight treatment teams will be randomly assigned to the test group and eight to the control group. The results of randomization will be made clear to the participating investigators and patients.

### Guarantee of independence of test and control groups

After randomization is completed, the treatment teams will strictly adhere to the assigned test or control protocol. In hospitals with treatment teams in both groups, patients in the treatment and control groups will be admitted to different wards to ensure no interference between them. An appointed hospital research supervisor will oversee protocol operations to ensure group independence.

### LTA testing

To ensure consistency and quality across different research centers, this study utilized an automatic platelet aggregator from the same manufacturer (AG800, Techlink Biomedical Technology Co., Ltd., Beijing, China). In each treatment team of test group, an automatic platelet aggregator was placed in the ward. Prior to the start of the study, the instrument was uniformly calibrated and debugged, using quality control products provided by the manufacturer for detection. Additionally, all participants underwent intensive training before being allowed to work on the sample collection and analysis process. This training included theoretical learning and the acquisition of operation skills, ensuring the standardization of the process.

The experimental conditions for the inspection process were standardized as follows:

(1) Specimen collection: After puncture, collect two 3ml tubes of fasting venous whole blood into citrate anticoagulated plastic vacuum blood collection tubes. (2) Plastic blood collection tubes were used for blood collection, and the plug containing magnesium ion was not used. (3) If the red blood cell specific volume (HCT) was greater than 55% during blood collection, the amount of anticoagulant was adjusted. (4) After blood collection (20∼25 ℃), the test was completed within 4 hours after storage, and the specimen was not vibrated during transportation. (5) All samples were collected between 8:00-10:00 a.m. and detected by computer within 4 hours after collection. (6) The platelet count of collected samples was greater than 100 × 109/L. (7) Centrifugation conditions for rich and poor plasma: platelet-rich plasma was centrifuged at 160 g / 10 min, and platelet-poor plasma was centrifuged at 2000 g / 10 min. (8) Inducer standards used: final concentration of ADP was 5 μmol / L, and the final concentration of arachidonic acid (AA) was 0.25 mg / ml. (9) Inducer storage conditions: after the powder dissolves into liquid, it can be stored at −20 ℃ for 1 month, and at 2∼8 ℃ for 1 week. (10) Instruments and consumables used: disposable cuvette (inner circle without dead angle and light transmittance), disposable mixing rod with two semicircular ends without acute angles, fixed stirring speed of 850 rpm, and a light source wavelength of 575 nm.

### Antiplatelet therapy and interventions

Prior to endovascular treatment, all patients will receive SDAT for at least 5 days. Patients in the test group will receive a guided antiplatelet regimen based on LTA testing; those with unsatisfactory platelet aggregation rates will undergo appropriate regimen adjustments (**Fig 2**). Test group patients will undergo drug adjustment at least 24 hours before stent implantation. LTA testing will be re-checked 48 hours after any drug regimen adjustment. All procedures will be performed under general anesthesia. Activated clotting time will be used to guide heparin administration during the procedure, with a target of 250 to 300 seconds. Typical dosing will be a 3000 to 5000 unit bolus at the start of the procedure and then 1000 units per hour. Stent selection will be based on anatomical measurements of the aneurysm and parent artery. After the procedure, patients will continue to take the same dual antiplatelet drugs as before.

**Figure 2.**
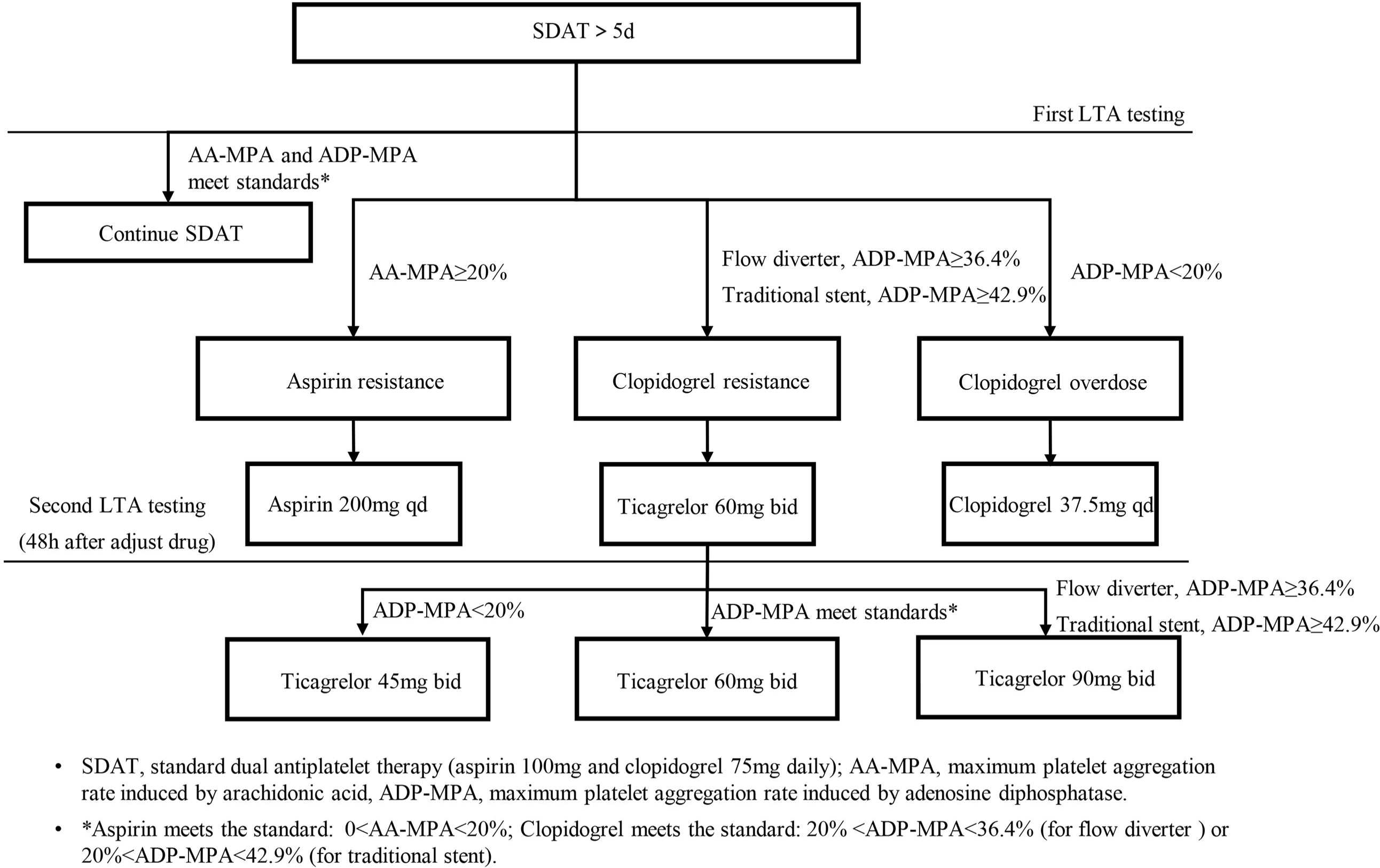
Light transmission aggregometry-guided antiplatelet therapy in the test group. SDAT, standard dual antiplatelet therapy (aspirin 100 mg and clopidogrel 75 mg daily); AA-MPA, maximum platelet aggregation rate induced by arachidonic acid, ADP-MPA, maximum platelet aggregation rate induced by adenosine diphosphate. *Aspirin meets the standard: 0<AA-MPA<20%; Clopidogrel meets the standard: 20% <ADP-MPA<36.4% (for flow diverter) or 20%<ADP-MPA<42.9% (for traditional stent).

Those who undergo placement of traditional stents will take clopidogrel or ticagrelor for at least 6 weeks and aspirin for at least 6 months. Patients who undergo flow diverter placement will take clopidogrel or ticagrelor for at least 3 months and aspirin for ≥2 years.

### Outcome measures

The primary outcome measure is ischemic events within the first 30 days after the procedure, including stent thrombosis, ischemic stroke, or transient ischemic attack. Stent thrombosis is defined as thrombosis at the stent site confirmed by digital subtraction angiography after stent implantation. Ischemic stroke is defined as rapid onset of a new focal neurological deficit with clinical or radiographic evidence of infarction, or rapid deterioration of an existing focal neurological deficit owing to a new infarction.^11^ Transient ischemic attack is defined as transient neurological dysfunction caused by focal cerebral ischemia.^12^

Exploratory outcome measures are ischemic events within the first 7 days of the procedure, modified Rankin scale score 30 days after the procedure and death from any cause.

The safety outcome is all bleeding events within 30 days of the procedure. Bleeding events will be graded according to the GUSTO grading criteria: 1) severe or life-threatening bleeding—intracranial bleeding or hemodynamically impaired bleeding requiring intervention; 2)moderate bleeding— bleeding requiring blood transfusion but not causing hemodynamic impairment; 3)minor bleeding—bleeding that does not meet the criteria for severe or moderate bleeding.^13^ Intracranial hemorrhage will be diagnosed using computed tomography or magnetic resonance imaging. Other types of hemorrhage will be diagnosed based on symptoms and imaging.

Study visits will be conducted on days 1, 3, 7, and 30 after the procedure to evaluate neurological status, ischemic events, and bleeding events. A five-member study committee will be formed to ensure safety and data reliability: a two-member data safety monitoring committee and three-member clinical event review committee. All ischemic and bleeding events will be assessed independently by each member of the clinical event committee. Committee members will be blinded to treatment group assignments. Any disagreements or discrepancies will be resolved via consensus.

### Subgroup analysis

The study will also assess whether guided antiplatelet therapy yields distinct therapeutic outcomes in various subgroups. Subgroup analyses will include a comparison between patients treated using flow diversion and those treated using traditional stent-assisted coil embolization, as well as stratified analyses based on factors such as age, sex, body mass index, smoking status, operation time, previous ischemic stroke, and the presence of combined atherosclerotic lesions.

### Sample size and data analysis

In a previous randomized controlled study of patients undergoing stent placement for intracranial aneurysm treatment, the incidence of ischemic events within 30 days of treatment in patients who received antiplatelet drug adjustment based on platelet function testing and those who received conventional treatment without monitoring was 5.1% and 12.1%, respectively.^14^ Based on a previous study, the intracluster correlation coefficient is assumed to be 0.002 in this study.^15^ With a test level of 0.05 and a power of 0.8, we anticipate recruiting 35 patients from each cluster (treatment team). Therefore, a total of 560 patients are required. Assuming a 5% attrition rate, 590 patients will be recruited (295 patients in the test and control groups, respectively). Each group will have one treatment team that recruits 36 patients; the other teams will recruit 37.

All analyses will be performed on the individual level with adjustment for clustering. All available data from the dropouts will be included in the analysis up to the time of dropout when possible. Statistical hypothesis tests will be two-tailed and performed at a 5% significance level. Continuous data will be tested for normality using the Shapiro–Wilk test. Normally distributed data will be expressed as means with standard deviation; skewed data will be expressed as medians with interquartile range. Categorical data will be expressed as numbers with percentage. Cluster-adjusted baseline differences between the two groups will be analyzed using the independent Student’s t test or Mann–Whitney U test for continuous variables and the Pearson χ2 test for categorical variables. All outcome event analyzes will be analyzed using generalized linear mixed-effects models with intervention as the fixed effect and the treatment team as the random effect to account for potential correlations in outcomes within each treatment team. Any variables that are unbalanced between the test and control groups (P <0.1) will be introduced into the model to adjust for confounders.

### Data collection and management

Researchers will enter data into the electronic data capture system in a timely fashion. All content required by the study plan in the system will be provided. Unfilled content will be explained. All outcomes arising during the study will be reported to the steering committee, which will review the reported events to ensure they meet the outcome definitions. Imaging data from computed tomography, magnetic resonance imaging, and digital subtraction angiography will be collected in DICOM format. Laboratory examination data will be collected by uploading photos. Imaging data will be sent to the designated data management center by the research inspector. The person in charge of the data management center will check and sign for receipt. All study files will be kept at all participating testing centers for at least 10 years. After study completion, the data will be reviewed by the research ethics committee, quality assurance committee, and other regulatory bodies. Study documentation in paper form (consent forms, questionnaires, and source datasets from the chart coordinator) will be securely stored. All computerized files will be password protected.

## DISCUSSION

In the neurointerventional treatment of intracranial aneurysms, SDAT is routinely administered to prevent thrombosis. However, the value of platelet function testing in patients receiving these drugs is controversial.^5,16–18^ A randomized controlled trial of coil embolization of unruptured cerebral aneurysms showed that modified antiplatelet therapy based on VerifyNow P2Y12 testing, which is commonly used because of its convenience, can reduce thromboembolic complications.^19^ However, LTA is the gold standard of platelet function testing and its value in patients undergoing endovascular interventional treatment of intracranial aneurysms has not yet been established. A single-center randomized controlled study confirmed that LTA-guided antiplatelet therapy in patients receiving ticagrelor helped reduce postoperative ischemic events; however, overall bleeding events significantly increased.^14^ Another study calculated optimal cut-off values for predicting ischemic events in patients who received traditional stents and flow diverters, which were 42.9% and 36.4% of the maximum platelet aggregation rates, respectively.^9^

Based on the above studies, we will conduct a multicenter prospective cluster randomized controlled study and adjust antiplatelet drugs according to the maximum platelet aggregation rate cut-offs. We hypothesize that LTA-guided antiplatelet therapy will reduce the incidence of ischemic events without increasing the incidence of major bleeding events compared with SDAT. The advantages of this study lie in the following aspects: multicenter design, standardized LTA testing across participating centers, customized cutoff values and antiplatelet regimens for flow diversion, and the use of low-dose ticagrelor. The results may provide evidence of a safe and effective alternative antiplatelet therapy for patients with intracranial aneurysms undergoing stent placement.

The study uses a cluster-randomized controlled design at the treatment team level in which the teams, not subjects, are randomized to avoid contamination of subjects in the same ward. Each treatment team only provides one type of treatment (test or control), which streamlines researcher training, alleviates the dilemma of physicians having to take both clinical treatment and research implementation into account, and avoids confusion.

We recognize the potential limitations of the trial. First, since the study will include only Asian patients, the results may not be applicable to other populations. Second, it focuses only on patients undergoing endovascular stenting for treatment of unruptured aneurysms. Therefore, the results will not apply to patients with ruptured aneurysms or those undergoing other stenting procedures. Third, because the study will be conducted in multiple centers, the LTA test results may be heterogeneous despite our best efforts to use the same testing equipment and uniform training protocols.

## Supporting information

trial protocol and statistical analysis plan

## Data Availability

All data produced in the present study are available upon reasonable request to the authors

