## Supplementary material for "Guided versus standard antiplatelet therapy in patients undergoing interventional intracranial aneurysm treatment using stents: protocol of a cluster randomized controlled cohort study": trial protocol and statistical analysis plan

**Sponsor:** Beijing Tiantan Hospital, Capital Medical University

**Principal Investigator:** Xinjian Yang, MD, Professor of Neurointervention

**Version No 1.1**

**Date: February 1, 2023**

#### Study site

| | |
|---|---|
| **Study title** | Guided versus standard antiplatelet therapy in patients undergoing interventional treatment of unruptured intracranial aneurysms using stents (GATITIA) |
| **Principal investigators (PIs)** | Professor Yang Xinjian<br>Beijing Tiantan Hospital, Capital Medical University<br>Address: No. 119, South 4th Ring Road West, Fengtai District, Beijing. |
| **Sponsor** | Beijing Tiantan Hospital, Capital Medical University |

#### Study outline

| | |
|---|---|
| **Study design** | Multicenter, parallel-arm, open-label, cluster-randomized, clinical trial |
| **Study objective** | To determine whether platelet function test-guided antiplatelet therapy can reduce the incidence of ischemic complications among patients undergoing endovascular intervention for intracranial aneurysms. |
| **Number of clusters** | 16 neurointerventional treatment teams planned |
| **Study outcome population** | Patients with unruptured intracranial aneurysms |
| **Primary endpoint** | Cerebral ischemic events within 30 days post-procedure [1-3] |
| **Exploratory outcome endpoints** | 1) Cerebral ischemic events within 7 days post-procedure;<br>2) Modified Rankin scale score at 30 days post-procedure;<br>3) All-cause mortality within 30 days post-procedure. |
| **Safety outcome endpoint** | All bleeding events within 30 days post-procedure. |

#### Neurointerventional treatment team inclusion criteria

| | |
|---|---|
| **Inclusion criteria** | 1) Team located in a hospital that can independently perform endovascular interventional therapy for intracranial aneurysms.<br>2) Attending interventionists on the team have >10 years of experience in aneurysm treatment.<br>3) The team performs an average of >10 interventional procedures each month for unruptured aneurysms. |

#### Participant inclusion and exclusion criteria

| | |
|---|---|
| **Inclusion criteria** | 1) Patients undergoing intracranial stenting for an unruptured intracranial aneurysm.<br>2) Standard dual antiplatelet therapy for ≥5 days before stenting.<br>3) Age 18–80 years.<br>4) Modified Rankin Scale score <2 on the day of enrollment.<br>5) Provision of written informed consent. |
| **Exclusion criteria** | 1) Recurrent aneurysm after interventional treatment or surgical clipping.<br>2) Allergy to aspirin, clopidogrel, or ticagrelor.<br>3) Prophylactic use of other antiplatelet drugs (e.g., tirofiban) before stent placement.<br>4) High probability of active bleeding because of symptomatic intracranial hemorrhage, active gastric ulcer, or history of bleeding or coagulation disorder.<br>5) Abnormal platelet count (normal range, 100–300 × $10^9$/L).<br>6) Ongoing use of anticoagulant medication.<br>7) Pregnancy or lactation.<br>8) Liver disease, kidney disease, congestive heart failure, or malignancy. |

#### Study Flow chart

| | Screening period | Enrollment period | Follow-up period | | |
|---|---|---|---|---|---|
| Follow-up time point | Before operation | Operation date | Post-operation | | |
| | - 14-0d | 0d | 3d | 7d | 30d±3d |
| | Visit 1 | Visit 2 | Visit 3 | Visit 4 | Visit 5 |
| Inclusion/exclusion criteria | X | | | | |
| Signing the ICF | X | | | | |
| Randomization | X | | | | |
| Demographic data | X | | | | |
| Medical history | X | | | | |
| Concomitant medication | X | X | X | X | X |
| Vital signs[1] | X | X | X | X | X |
| mRS | X | X | X | X | X |
| Blood routine[2], blood Lipids[3], blood bio-Chemistry[4], Coagulation[5] | X | | | | X |
| Cerebrovascular imaging examination | X | X | | | |
| Platelet function testing [6] | X | | | | |
| Drug distribution/recovery[7] | X | X | X | X | X |
| Investigation of patient compliance | X | X | X | X | X |
| Medication prescription | X | X | X | X | X |
| AE, SAE and Endpoint event[8] | | X | X | X | X |

Notes:

1) Vital signs: Heart rate, blood pressure, body temperature and respiration rate;
2) Blood routine: Red blood cell count, white blood cell count, platelet count and hemoglobin;
3) Blood lipids: Total cholesterol, triglyceride, high-density lipoprotein cholesterol, low-density lipoprotein cholesterol;
4) Blood biochemistry: Alanine aminotransferase, aspartate aminotransferase, alkaline phosphatase, Gamma-glutamyl transferase, total bilirubin, blood urea nitrogen, blood creatinine;
5) Coagulation function: Prothrombin time, fibrinogen, activated partial thromboplastin time, thrombin time, international normalized ratio (INR);
6) Platelet function testing is platelet aggregation rate test;
7) Drug distribution/return, compliance and medication prescription are evaluated for the use of aspirin, clopidogrel and ticagrelor.
8) Adverse events (AE), serious adverse events (SAE) and endpoint events are the occurrence of AE, SAE and endpoint events of concern during the operation period and follow-up period.

#### Abbreviations

| Abbreviations | Interpretation of the meaning |
| --- | --- |
| AA | Arachidonic acid |
| AA-MPA | Maximum platelet aggregation rate induced by arachidonic acid |
| ADP | Adenosine 5′-diphosphate |
| ADP-MPA | Maximum platelet aggregation rate induced by adenosine diphosphate |
| AE | Adverse event |
| CI | Confidence interval |
| CYP2C19 | Cytochrome P450 2C19 |
| CYP3A | Cytochrome P450 3A |
| EDC | Electronic Data Capture |
| GCP | Good clinical practice |
| GUSTO | Global Use of Strategies to Open Occluded Coronary Arteries |
| HTPR | High on-treatment platelet reactivity |
| ICF | Informed consent forms |
| ITT | Intention to treat |
| LTA | Light transmittance aggregometry |
| LTPR | Low on-treatment platelet reactivity |
| MPA | Maximal aggregation |
| mRS | Modified Rankin Scale |
| SAE | Serious adverse event |
| SDAT | Standard dual antiplatelet therapy |

#### 1. Study Background

Intracranial aneurysms are generally treated by endovascular interventional therapy when possible.[4,5] The use of intracranial stents, especially flow diverters, has considerably expanded the indications for interventional therapy and contributed to higher treatment success rates. However, the presence of an endovascular stent also increases the risk of ischemic complications.[6,7] Standard dual antiplatelet therapy (SDAT), consisting of oral aspirin 100 mg and clopidogrel 75 mg daily, is administered to patients undergoing stent placement to reduce the incidence of thrombotic events. However, the response to antiplatelet drugs varies among patients. Some patients develop high on-treatment platelet reactivity (HTPR) to antiplatelet drugs, such as clopidogrel, and the incidence of thrombotic events can reach 40%.[8]

Platelet function tests are important for identifying patients with resistance to antiplatelet drugs. The escalation of antiplatelet therapy in such patients may reduce the risk of ischemic complications.[9,10] Light transmittance aggregometry (LTA) is the gold standard for evaluating platelet aggregation function, which may be important in patients undergoing interventional treatment for intracranial aneurysms. The maximum platelet aggregation rate induced by adenosine diphosphate (ADP-MPA) can be used to predict ischemic events on the basis of previously established cutoff values.[11]

Ticagrelor is a reversible P2Y12 receptor antagonist with a rapid onset of action. Unlike clopidogrel, its activity does not require conversion by CYP450 enzymes, and its efficacy is not affected by CYP2C19 gene variants.[12] Therefore, we will conduct a cluster-randomized controlled study to determine whether LTA-guided antiplatelet therapy can reduce the incidence of ischemic complications among patients undergoing endovascular intervention for unruptured intracranial aneurysms.

#### 2. Study Objective

To determine whether platelet function test-guided antiplatelet therapy can reduce the incidence of ischemic complications, compared with SDAT, among patients undergoing endovascular intervention for unruptured intracranial aneurysms.

#### 3. Study Design

This prospective, multicenter, open-label, cluster-randomized controlled study with blinded outcome adjudication will include 16 neurointerventional treatment teams from eight hospitals. The 16 teams will be randomly divided into test and control groups at a ratio of 1:1. Patients in the test group will receive LTA test-guided antiplatelet therapy, whereas control group patients will receive SDAT throughout the study period.

#### 4. Study Outcomes

##### 4.1 Primary study outcomes

The primary outcome measure is a composite of ischemic events within the first 30 days post-procedure, including stent thrombosis, ischemic stroke, and transient ischemic attack. Stent thrombosis is defined as thrombosis at the stent site or distal cerebral vessels confirmed by digital subtraction angiography after stent implantation. Ischemic stroke is defined as rapid onset of a new focal neurological deficit with clinical or radiographic evidence of infarction, or rapid deterioration of an existing focal neurological deficit caused by a new infarction.[3] Transient ischemic attack is defined as transient neurological dysfunction caused by focal cerebral ischemia.[2] Details of the diagnostic criteria for ischemic events are presented in Appendix 1.

##### 4.2 Exploratory study outcomes

Exploratory outcome measures are ischemic events within the first 7 days post-procedure, modified Rankin scale (mRS) score at 30 days post-procedure, and all-cause mortality within 30 days post-procedure.

**4.3 Safety study outcome**

The safety outcome is all bleeding events within 30 days post-procedure. Bleeding events will be graded according to the Global Utilization of Streptokinase and Tissue Plasminogen Activator for Occluded Coronary Arteries (GUSTO) criteria: 1) severe or life-threatening bleeding—intracranial bleeding or hemodynamically impaired bleeding requiring intervention; 2) moderate bleeding—bleeding requiring blood transfusion but not causing hemodynamic impairment; 3) minor bleeding—bleeding that does not meet the criteria for severe or moderate bleeding.[13]

**4.4 Study outcome evaluation**

A five-member study committee will be formed to ensure safety and data reliability. This committee will consist of a two-member data safety monitoring committee and a three-member clinical events committee. All primary outcomes, exploratory outcomes, and safety outcomes will be independently adjudicated by three members of the clinical events committee. A third member of the clinical events committee will resolve disagreements. Discrepancies will be reviewed by all five members of the committee and resolved by consensus. Imaging results related to clinical events will be evaluated at an independent imaging core laboratory by trained staff who are blinded to treatment group assignments. The above-mentioned committee members will also be blinded to treatment group assignments.

#### 5. Study clusters

In this study, a treatment team is defined as a cluster containing ≥1 neurointerventional expert skilled in neurointerventional treatment for aneurysms, one attending physician proficient in perioperative management of aneurysms, and one caregiver experienced in neurointerventional surgical care. The treatment team selection criteria are as follows.

1) The team is located in a hospital that can independently perform endovascular interventional therapy for intracranial aneurysms.

2) Attending interventionists on the team have >10 years of experience in aneurysm treatment.

3) The team performs an average of >10 interventional procedures each month for unruptured aneurysms.

#### 6. Study Population

##### 6.1 Inclusion criteria

1) Patients undergoing intracranial stenting for an unruptured intracranial aneurysm.

2) SDAT for ≥5 days before stenting.

3) Age 18–80 years.

4) Modified Rankin Scale score <2 on the day of enrollment.

5) Provision of informed consent form (ICF).

##### 6.2 Exclusion criteria

1) Recurrent aneurysm after interventional treatment or surgical clipping.

2) Allergy to aspirin, clopidogrel, or ticagrelor.

3) Prophylactic use of other antiplatelet drugs (e.g., tirofiban) before stent placement.

4) High probability of active bleeding because of symptomatic intracranial hemorrhage, active gastric ulcer, or history of bleeding or coagulation disorder.

5) Abnormal platelet count (normal range, 100–300 × $10^9$/L).

6) Ongoing use of anticoagulant medication.

7) Pregnancy or lactation.

8) Liver disease, kidney disease, congestive heart failure, or malignancy.

#### 7. Treatment of participants

##### 7.1 Antiplatelet drugs

Aspirin functions by suppressing the production of prostaglandins and thromboxane through irreversible inactivation of cyclooxygenase, which is necessary for the synthesis of these factors. Low doses of aspirin block the formation of thromboxane A2 (an arachidonic acid metabolite) in platelets, thereby inhibiting platelet aggregation throughout the lifespan of the affected platelets. Clopidogrel, a prodrug requiring CYP2C19 for activation, acts on the adenosine diphosphate (ADP) receptor present on platelet cell membranes; it specifically and irreversibly inhibits the P2Y12 subtype of the ADP receptor. This subtype of the ADP receptor is essential for platelet activation and eventual cross-linking by fibrin. Finally, ticagrelor is a reversible and direct-acting oral antagonist of the adenosine diphosphate receptor P2Y12; it provides more rapid, stronger, and consistent P2Y12 inhibition compared with clopidogrel.

##### 7.2 Platelet function test

###### 7.2.1 Patient preparation.

Ideally, patients should rest before blood collection to reduce the effects of exercise-induced epinephrine release on platelet aggregation. Patients should stop smoking for ≥30 minutes, avoid caffeine for ≥2 hours, and avoid eating greasy foods on the day of blood draw.

###### 7.2.2 Blood sampling.

Blood samples for platelet function analyses are collected from an antecubital vein using a 21-gauge needle, 2–4 h after antiplatelet drug intake. The first 2–4 mL of blood are discarded to avoid spontaneous platelet activation, and samples are processed within 1 hour.

###### 7.2.3 Platelet function testing

Platelet function testing involves assessment of platelet aggregation through LTA and evaluation of platelet activation markers via flow cytometry. To determine platelet reactivity to clopidogrel and aspirin, we will measure the platelet aggregation rates induced by adenosine diphosphate (ADP-MPA) and arachidonic acid (AA-MPA), respectively. To ensure consistency and quality across research centers, we will utilize an automatic platelet aggregator from the same manufacturer (AG800, Techlink Biomedical Technology Co., Ltd., Beijing, China). An automatic platelet aggregator will be placed in the ward of each treatment team in the test group. Prior to the start of the study, the instrument will be uniformly calibrated and debugged using quality control products provided by the manufacturer. Additionally, all participating teams will undergo intensive training before performing sample collection and analysis procedures. This training will include theoretical learning and the acquisition of technical skills to ensure standardized testing.

The standardized experimental conditions for the inspection process are as follows:

1) Specimen collection: After puncture, collect two 3-ml tubes of fasting venous whole blood into citrate anticoagulated plastic vacuum blood collection tubes.

2) Plastic blood collection tubes are used for blood collection; no magnesium ion-containing plugs are used.

3) If a red blood cell-specific volume exceeds 55% during blood collection, the amount of anticoagulant is adjusted.

4) After blood collection (20–25°C), the test is completed within 4 hours, and the specimen is not shaken during transportation.

5) All samples are collected between 8:00 and 10:00 a.m.

6) The platelet counts of collected samples are >100 × $10^9$/L.

7) Centrifuge conditions for platelet-rich and platelet-poor plasma: platelet-rich plasma is centrifuged at 160 x g for 10 minutes, and platelet-poor

plasma is centrifuged at 2000 x g for 10 minutes.

8) Inducer standards used: the final concentration of ADP is 5 μmol/L, and the final concentration of arachidonic acid (AA) is 0.25 mg/ml.

9) Inducer storage conditions: after the powder has been dissolved in liquid, it can be stored at -20°C for 1 month and at 2–8°C for 1 week.

10) Instruments and consumables used: disposable cuvette (inner ring lacking dead angles and light transmittance), disposable mixing rod with two semicircular ends without acute angles, fixed stirring speed of 850 rpm, and light source wavelength of 575 nm.

11) Platelet aggregation curves are recorded for 5 minutes and maximal aggregation (MPA) is measured in relation to the highest percentage of stimulus-induced platelet aggregation.

**7.2.4 Definitions of aspirin and clopidogrel responses**

Our previous study showed that the cutoff values for predicting ischemic events differ between traditional stents and flow diverters.[11] Therefore, this study uses stent-specific HTPR cutoff values. For traditional stents, HTPR to clopidogrel is regarded as ADP-MPA ≥42.9%; for flow diverters, HTPR to clopidogrel is regarded as ADP-MPA ≥36.4%. However, no definitive studies have confirmed a specific threshold for platelet reactivity to AA that is associated with thrombotic risk among patients with intracranial aneurysms after stent placement. Based on insights from a cardiology trial, aspirin resistance is defined as AA-MPA ≥20%.[14] Additionally, a low on-treatment platelet reactivity (LTPR) response to ADP is defined as ADP-MPA <20% and AA-MPA=0%.

**7.3 Antiplatelet therapy regimen**

All patients will receive clopidogrel (75 mg/day) and aspirin (100 mg/day) for ≥5 days before stent implantation. Patients in the control group will receive SDAT throughout the study period. PFT will not be performed on patients in the control group, mainly for the following reasons. Firstly, as per the previous study design.[15,16] Secondly, from a statistical standpoint, baselines are comparable when conducting random assignment. Thirdly, this study is an open-label study, and the lack of LTA testing in the control group helps avoid interference to clinicians. Lastly, whether to perform routine platelet aggregation function testing before neurointerventional procedure remains a controversial topic. A design plan was adopted in which the platelet function was not tested in the control group. The final comparison of results will answer this important clinical question.

Patients in the test group will undergo LTA testing at 5 days after initiation of SDAT. Patients with HTPR or LTPR will undergo antiplatelet drug adjustment. Patients with HTPR to aspirin will receive aspirin 200 mg once daily, patients with HTPR to clopidogrel will receive ticagrelor 60 mg twice daily, and patients with LTPR to clopidogrel will receive clopidogrel 37.5 mg once daily. A second LTA test will be performed 48 hours after adjustment. Patients who exhibit HTPR to ticagrelor will receive ticagrelor 90 mg twice daily. Patients who exhibit LTPR to ticagrelor will receive ticagrelor 45 mg twice daily. Details of the antiplatelet regimen for patients in the test group are presented in Appendix 4.

**7.4 Routine protocol for stenting treatment of patients with unruptured intracranial aneurysm**

Stent placement will occur 24 hours after drug adjustment in the test group. All procedures will be performed under general anesthesia. Systemic heparinization will be performed after the placement of a femoral introducer sheath. Prior to embolization, rotational angiography will be conducted, followed by three-dimensional image reconstruction through volume rendering. Based on images generated via rotational acquisition, ≥2 working projections that provide the best view of the aneurysm neck will be defined. Stent-assisted coiling will be used for aneurysms that cannot undergo standard coiling. Properly shaped microcatheters will be introduced over micro guidewires and navigated into the aneurysm cavity under the guidance of the micro guidewire. During the coiling procedure, a coil suitable for safe packing will be selected at each step. Aneurysms will be packed as densely as possible with coils. The jailing technique, where a stent is placed after the microcatheter is in position and the first coil is placed but not detached, will typically be used. For complex intracranial aneurysms (e.g., large or giant, fusiform, etc.), a flow diverter will be selected for complete occlusion.[7] A triaxial support system will be implemented to access aneurysms in patients receiving flow diverters. The flow diverter will be introduced through a microcatheter, delivered to the parent artery defect, and then placed. Several endovascular techniques (e.g., use of wires, catheters, or balloon angioplasty) will be performed if the device is inadequately expanded. Similar techniques, including stent-assisted coiling, will be implemented for aneurysms treated with flow diverters and coiling. A final postembolization angiography will be conducted in the working projection to assess the occlusion grade and detect any complications. Frontal and lateral projections will be acquired at the end of the procedure.

##### 7.5 Postoperative management

After the procedure, patients will be observed in a recovery unit for 30–60 minutes. If the procedure is successful and the patient fully recovers from general anesthesia without any complications, they will be transferred to a general ward. Over the next 24 hours, attending nurses will monitor the patient in a routine manner. During the daytime, physician assistants will check on the patient every 2 hours; during the nighttime, on-duty residents will perform checks every 4 hours. If any events occur, the attending nurse will notify the physician assistants, on-duty residents, or operating neurointerventionists, who will assess the patient's status. In cases where patients exhibit symptoms of neurological deficit post-procedure, distinguishing between a bleeding event and an ischemic event can be challenging within a short time. Due to its quicker completion time, a CT scan is typically the initial choice to exclude cerebral hemorrhage. If a CT scan reveals brain hemorrhage, an MRI scan is unnecessary. However, if no evidence of brain hemorrhage is found on the CT scan, an MRI scan becomes necessary. In summary, MRI scans were conducted on all patients with cerebral infarction who presented with symptoms of neurological deficit. In cases of transient ischemic attack and stroke, the critical pathway for in-hospital stroke will be activated. If there is evidence of thrombus or embolus causing flow disturbance, intra-arterial fibrinolytics or antiplatelet drugs (e.g., tirofiban or abciximab) will be used. If necessary, conventional angiography and intervention may be included for evaluation and management. For other cases, such as bleeding complications, appropriate management will be determined based on case severity. Patients who do not experience any events during the first 48 hours after stenting will be discharged. Ischemic events will be evaluated through clinical examination or imaging. After discharge, patients will be advised to visit our hospital, regardless of an appointment, if any problems related to the stent placement arise after receiving medical care at a primary or secondary care center near their home.

**7.6 Follow-up management**

After the procedure, patients will continue taking the same dual antiplatelet drugs they received before the procedure. Patients who undergo placement of traditional stents will receive clopidogrel or ticagrelor for ≥6 weeks and aspirin for ≥6 months. Patients who undergo flow diverter placement will receive clopidogrel or ticagrelor for ≥3 months and aspirin for ≥2 years.[17] Angiographic follow-up studies will be routinely performed in patients at 6 months after treatment; if the results are stable, additional follow-up angiographs will be conducted over longer intervals (e.g., annually). Patients with a potential risk of recanalization will undergo repeated angiographs. Clinical follow-up results will be assessed during hospitalization for follow-up angiography or through a telephone interview. Any ischemic events within 7 days and 1 month, as well as bleeding events within 1 month, will be recorded.

#### 8. Neurointerventional product

| Device name | Manufacturer | Description |
|---|---|---|
| Solitaire | Medtronic | Traditional Stent |
| Neuroform EZ | Stryker | Traditional Stent |
| Atalas | Stryker | Traditional Stent |
| Enterprise stent | Codman | Traditional Stent |
| Low-profile Visualized Intraluminal Support stent | MicroVention | Traditional Stent |
| Pipeline embolization device | ev3 | Flow diverter |
| Tubridge flow diverter | MicroPort | Flow diverter |
| Surpass Streamline | Stryker | Flow diverter |
| Target series | Stryker | Coils |
| Axium™ series | Medtronic | Coils |
| Galaxy series | Codman | Coils |
| Avenir™ | Wallaby Medical | Coils |
| Jasper, Presgo | Jiaqi Shengwu | Coils |
| Visee | Visee Medical | Coils |
| Perdenser, Perfiller | Lepu Medical Technology | Coils |

#### 9 Concomitant Medication

During the study period, any new medications, surgeries, and changes to medications will be recorded.

##### 9.1 Prohibited concomitant treatments

1) Antiplatelet agents: dipyridamole, cilostazol, ticlopidine, prasugrel, GPIIb/IIIa receptor antagonist (tirofiban is allowed for thrombosis), ozagrel, and similar agents.

2) All anticoagulants.

3) Thrombolytic drugs: rt-PA, urokinase, streptokinase, and similar agents.

4) Batroxobin, snake venom preparation, lumbrokinase, and similar agents.

5) Potent CYP3A inhibitors: ketoconazole, nefazodone, ritonavir, saquinavir, atazanavir, nelfinavir, itraconazole, voriconazole, clarithromycin, telithromycin (excluding erythromycin or azithromycin).

6)Potent CYP3A inducers: rifampicin, dexamethasone, phenytoin, carbamazepine, oxcarbazepine, and phenobarbital.

7) Potent CYP2C19 inhibitors: omeprazole, esomeprazole; fluvoxamine, fluoxetine, moclobemide, fluconazole, ciprofloxacin and cimetidine.

8) Potent P-glycoprotein inhibitors: verapamil, quinidine, and cyclosporine.

9) P-glycoprotein substrate: digoxin. If digoxin is absolutely necessary, then the dose of digoxin will be reduced as appropriate and the blood concentration of digoxin will be closely monitored.

If use of the above drugs violates the protocol, study enrollment will be terminated, and the reasons will be recorded; however, the visit on day 30 ± 3 should be completed.

**9.2 Permitted concomitant treatments**

Any drugs other than those listed above are permitted. Patients with underlying diseases (e.g., hypertension, diabetes, coronary heart disease, and epilepsy) can use diuretics, beta blockers, angiotensin-converting enzyme inhibitors, angiotensin receptor blockers, calcium antagonists, lipid-lowering medications, coronary vasodilators, anti-diabetic medications (including insulin), anti-epileptics, and other necessary agents. Information about diagnosis, medication, dose, and administration will be collected. Any required treatments for concomitant diseases during the study period must be recorded, including information about diagnosis, medication, dose, and administration.

1) H2 receptor blockers are permitted (except cimetidine).

2) Proton pump inhibitors: Except for omeprazole and esomeprazole, other types of proton pump inhibitors can be used. Rabeprazole is recommended. Other proton pump inhibitors can be used, such as dexlansoprazole, lansoprazole, and pantoprazole.
3) Statin: doses of simvastatin or lovastatin ≤40 mg daily or any dose of any other statin will be permitted.
4) For drugs that can induce bradycardia (e.g., beta blockers, and calcium antagonists) when combined with ticagrelor, heart rate will be closely monitored.
5) Serotonin reuptake inhibitors (e.g., paroxetine, sertraline, and citalopram), which may increase the risk of bleeding. If the concomitant use of serotonin reuptake inhibitors is inevitable, bleeding events will be carefully monitored.
For any drugs used during study, the generic name (non-trade name), dosage and administration method, reason for use, and start and end time of use will be recorded in detail in the electronic data capture (EDC) system.

#### 10. Visit Procedures

A trained clinical research coordinator who is unaware of the treatment allocation must record the patient’s routine visit data and collect the information and data required in the clinical study flowchart.

##### 10.1 Visit 1 (-14 to 0 days before operation)

All patients who meet all inclusion criteria and do not meet any exclusion criteria and voluntarily sign an ICF will be asked to undergo imaging evaluation and complete the following tasks.

1) Provision of the written ICF.

2) Verification of inclusion and exclusion criteria.

3) Documentation of age, sex, height, weight, body mass index, alcohol intake history, smoking history, and ethnicity.

4) Documentation of the time and course of cerebral ischemic symptom events.

5) Documentation of medical history (e.g., hypertension, diabetes, coronary heart disease, and hyperlipidemia).

6) Documentation of concomitant medications (type, dose, and frequency).

7) Documentation of vital signs (blood pressure, heart rate, respiration rate, and pulse).

8) Physical examination (focusing on stroke-relate contents) and mRS assessment.

9) Collection of fasting blood samples and performance of routine blood tests (red blood cell count, white blood cell count, platelet count, and hemoglobin), blood lipid assessments (total cholesterol, triglycerides, high-density lipoprotein cholesterol, and low-density lipoprotein cholesterol), blood biochemistry analysis (alanine amino acid transferase, aspartate aminotransferase, alkaline phosphatase, γ-glutamyl transferase, total bilirubin, blood urea nitrogen, and blood creatinine), and coagulation function evaluation (e.g., prothrombin time, fibrinogen, activation Partial thromboplastin time, and thrombin time).

10) Digital subtraction angiography assessment of aneurysm characteristics—size, number, location, shape, neck width, and diameter of parent artery.

All patients recruited into the test group must undergo LTA testing before procedure, and antiplatelet drugs will be adjusted based on the test results. The following information should be recorded:

11) LTA test results, including AA-MPA and ADP-MPA.

12) Antiplatelet adjustment regimen.

13) Patient compliance.

**10.2 Visit 2 (Date of operation)**

This visit occurs during the operation, and the details of operation should be recorded. The following tasks should be completed at this visit:

1) Documentation of vital signs: heart rate, blood pressure, body temperature, and respiration rate before operation.

2) Aneurysm characteristics—size, number, location, shape, neck width, and diameter of parent artery.

2) Stents and coils (name, manufacturer, quantity, and specification).

3) Aneurysm embolization results.

4) Documentation of concomitant medication.

5) Digital subtraction angiography assessment of cerebral vessels.

6) Documentation of any adverse events (e.g., ischemic events, bleeding events, or death).

7) mRS assessment.

8) Scheduling the date and time of next visit.

**10.3 Visit 3 (3 days after stent implantation)**

Visit 3 will be performed at 3 days post-procedure. The following tasks should be completed at this visit:

1) Documentation of antiplatelet treatment regimen.

2) Documentation of concomitant medication.

3) Documentation of vital signs: heart rate, blood pressure, body temperature, and respiration rate.

4) Documentation of any adverse events (e.g., ischemic events, bleeding events, or death).

5) mRS assessment.

6) Scheduling the date and time of next visit.

**10.4 Visit 4 (7 days after stent implantation)**

Visit 4 will be performed at 7 days post-procedure. The following tasks should be completed at this visit:

1) Documentation of antiplatelet treatment regimen.

2) Documentation of concomitant medication.

3) Documentation of vital signs: heart rate, blood pressure, body temperature, and respiration rate.

4) Documentation of any adverse events (e.g., ischemic events, bleeding events, or death).

5) mRS assessment.

6) Scheduling the date and time of next visit.

**10.5 Visit 5 (30 ± 3days after stent implantation)**

Visit 5 will be performed at 30 days post-procedure. The following tasks should be completed at this visit:

1) Documentation of antiplatelet treatment regimen.

2) Documentation of concomitant medication.

3) Documentation of vital signs: heart rate, blood pressure, body temperature and respiration rate.

4) Documentation of any adverse events (e.g., ischemic events, bleeding events, or death).

5) mRS assessment.

6) Collection of fasting blood samples and performance of routine blood tests, assessment of blood lipids, blood biochemistry analysis, and coagulation function evaluation.

#### 11. Safety Reporting

##### 11.1 Definitions of adverse events

Any adverse medical event that occurs from patient randomization to the end of follow-up, regardless of whether its causal relationship with the antiplatelet drug, is considered an adverse event. Any event that occurs in a participant in a clinical study that is not expected to occur is also an adverse event. Research personnel will report all adverse events directly observed by clinical staff or spontaneously reported by participants in concise medical terminology. Adverse drug reactions refer to reactions directly or indirectly related to drugs. To prevent omission, if there is difficulty in determining the relationship to the drug, any adverse event that occurs should be recorded, documenting the time of occurrence, severity, duration, measures taken, and regression of the adverse event, as well as its possible relationship with the study drug.

**11.2 Observation and documentation of adverse events**

A safety assessment will be conducted on all participants; all events including adverse events (AEs), severe adverse events (SAEs), death, ischemic event, bleeding event, discontinuation of medication for any reason, abnormal laboratory tests, changes in vital signs, and changes in physical examination results will be recorded. Adverse events will be recorded from randomization until 30 days after operation. If an adverse event occurs, it will be monitored until the adverse event disappears, returns to baseline levels, or is confirmed as not clinically significant. Evaluations of adverse events will include the name and severity of each event, regression information, relationship with antiplatelet drugs, and management measures.

**11.3 Severities of adverse events**

The severities of adverse events will be divided into three levels: mild, moderate, and severe, defined as follows.

1) Mild: Mild symptoms or illnesses that rapidly improve after discontinuation of medication without requiring treatment.

2) Moderate: Temporary damage not requiring hospitalization or extended hospitalization, requiring treatment or intervention, easily recovered.

3) Severe: Events causing transient damage, requiring hospitalization for outpatients and extended hospitalization for inpatients (>7 days), causing permanent system/organ damage, or considered life-threatening (e.g., symptoms requiring emergency care such as asphyxia, shock, or coma).

**11.4 Correlations between adverse events and antiplatelet drugs**

The investigator should evaluate the potential link between the adverse event and the antiplatelet drug using a 5-level classification system.

1) Definitely related: The adverse reaction occurs in a logical sequence following drug administration, aligns with known reactions attributed to the drug, improves upon discontinuation, and reoccurs with re-administration. This reaction cannot be attributed to the participant's underlying condition or the use of other medications.

2) Likely related: The reaction emerges in a sensible chronological order after the drug is given, matches the expected reaction profile for the drug, improves when the drug is stopped, and cannot be attributed to the participant’s existing health issues or the use of other medications.

3) Possibly related: The reaction appears within a plausible timeframe after drug administration, is consistent with the drug's known reaction profile, but the patient's clinical status or another treatment could also contribute to the reaction.

4) Possibly unrelated: The reaction does not occur within a reasonable timeframe after drug administration, nor does it match the expected reaction profile of the drug, suggesting the patient's health status or another treatment could be the cause.

5) Unrelated: The reaction does not follow a logical timeframe after administration, fits the reaction profile of a different drug, and is likely caused by the patient's health condition or another treatment. Improvement or resolution occurs when other treatments are stopped, and the reaction reoccurs with the reintroduction of these treatments.

##### 11.5 Serious Adverse Events

Serious adverse events are defined as:

1) Causing death.

2) Life-threatening (this refers to the risk of death at the time of the event and not to events that could potentially lead to death if conditions worsen).

3) Requiring hospitalization or prolonging hospital stay.

4) Resulting in significant or persistent disability or dysfunction.

5) Leading to congenital anomalies or birth defects.

6) Considered by researchers to be a serious medical incident.

Should a serious adverse event occur during the trial, irrespective of its relation to the drug, the investigator must immediately implement appropriate treatment measures for the participant’s safety and report the event to the ethics committee at the center. Additionally, completion of a serious adverse event form is required. Any serious adverse event that is unresolved by the trial's conclusion or upon the participant’s early withdrawal should be monitored until one of the following conditions is met:

1) The event disappears or is resolved.

2) Events are stable.

3) Return the event to the baseline level (if baseline values are available).

4) The event was relieved to no clinical significance.

5) The event can be attributed to a medication other than the study drug or factors unrelated to the conduct of the study, or when it is not possible to provide more information.

**11.6 Recording, Processing, and Reporting**

*****All adverse events, whether related to the antiplatelet drug or not, must be carefully documented.

*****Handling drug-related adverse events meticulously is crucial.

*****Participants should be encouraged to truthfully report any changes in their condition after taking the drug.

*****Researchers should avoid leading questions and closely monitor for any adverse events or unexpected side effects, analyzing causes, making judgments, and documenting follow-up observations.

*****The incidence of adverse events should be recorded.

*****Detailed information on adverse events during the trial, including symptoms, severity, onset, duration, treatment actions, outcomes, drug relationship, and follow-up strategies, should be recorded.

*In cases of adverse events, the researcher may suspend the study to conduct detailed investigations on discontinued cases, documenting the treatment and outcomes.

*In the event of a serious or important adverse event, emergency procedures must be initiated by the investigator.

*All trial participants who have used the therapeutic drug should be included in the adverse event statistics, regardless of trial completion.

##### 11.7 Emergency Procedures

Upon occurrence of a serious or significant adverse event, researchers must immediately organize rescue efforts to ensure participant safety, notifying the responsible unit via phone/fax regardless of the relation to the antiplatelet drug. The research center folder contains detailed reporting procedures. A written report detailing the event's circumstances and outcomes must follow the initial notification, complementing the phone/fax report with additional information if necessary. Clinical monitoring staff should review and receive written reports within 24 hours or by the second working day, following Good Clinical Practice (GCP) guidelines.

#### 12 Data Recording and Management

##### 12.1 Data Entry and Confidentiality

An Electronic Data Capture (EDC) system is utilized for data collection. Data are prospectively collected by research coordinators who are not involved in patient care, across all participating treatment teams. These coordinators are compensated for their participation in the trial. Prior to the official launch of the EDC system, relevant users will undergo training and testing to ensure the system fulfills the study's requirements. Upon launch, research coordinators will receive their account details, including a username and password. Each account is specifically tied to the user's role and permissions. It is imperative that account information is securely managed and not shared with others, nor should anyone exercise rights on behalf of another user. The EDC system facilitates direct data transfer from the client to the server over the internet. Data collection is achieved by directly inputting source data into the EDC system. Research coordinators are tasked with ensuring the quality of the input data to maintain its authenticity and integrity. The system also features an interface printing function, allowing for the printing of electronic case report form information as needed. Confidentiality of participants' personal data is paramount; although the ethics committee and implementers may access participant information, they are prohibited from disclosing any details. All researchers are obligated to keep trial results, plans, and materials strictly confidential, with information release contingent upon written authorization from the principal investigator. Research implementation, protocols, design, results, data collection, and any related materials are to be confidentially maintained by the project's researchers, accessible only to authorized individuals.

##### 12.2 Data locking

After data review and confirmation of database accuracy, the principal investigator and statistical analyst will lock the data. Once locked, the data or documents will no longer be subject to changes. Any issues identified post-lock will be addressed in the statistical analysis program.

##### 12.3 Data processing

Upon database locking, the database is forwarded to the statistical analyst for analysis as per the statistical analysis plan. Following the completion of statistical analysis, the analyst will draft a statistical analysis report and submit it to the trial's principal investigator for the development of the clinical study report.

#### 13 Statistics and Data Analysis

Statistical analysis for the study will be conducted by a third party, using SPSS software version 26.0 (IBM Corp., Armonk, NY, USA).

##### 13.1 Calculation of sample size

Based on literature results for patients undergoing stent placement for intracranial aneurysm treatment, it is assumed that the incidence of ischemic events within 30 days of treatment is 5.1% for patients receiving antiplatelet drug adjustment based on platelet function testing and 12.1% for those receiving standard drug therapy without monitoring.[18] Assuming an intracluster correlation coefficient of 0.002,[19] with a test level of 0.05 and a power of 0.8, it is anticipated that 35 patients will be recruited from each cluster (treatment team), requiring a total of 560 patients. Considering a 5% attrition rate, the recruitment target is set at 590 patients (295 per group).

##### 13.2 Statistical method

The primary analysis will use an intention-to-treat (ITT) protocol. All participants who underwent random grouping and completed stent placement constitute the full analysis set for this study. The full analysis set is used to analyze baseline information and endpoint indicators; it will be regarded as the dataset for endpoint evaluation in this study. Missing values will be left as is, with patients censored at their last follow-up. Statistical hypothesis tests will be two-tailed, performed at a 5% significance level.

###### 13.2.1 Balance of baseline characteristics

Normality of continuous data will be tested using the Shapiro–Wilk test. Normally distributed data will be presented as means with standard deviation; skewed data as medians with interquartile range. Categorical data will be presented as numbers with percentages. Differences at baseline between groups will be analyzed using the independent Student's t-test or Mann–Whitney U test for continuous variables and the Pearson $\chi 2$ test for categorical variables.

##### 13.2.2 Primary efficacy measures

The rate of ischemic events (composite of stent thrombosis, ischemic stroke, and transient ischemic attack) during the 30-day treatment period for the ITT population will be summarized. Primary endpoint analysis will use generalized linear mixed-effects models, with intervention as the fixed effect and treatment team as the random effect, to account for correlations within treatment teams. Variables significantly unbalanced between groups (P<.1) will be included in the model to adjust for confounders.

##### 13.2.3 Exploratory efficacy measures:

The rate of ischemic events during the 7-day treatment period for the ITT population, along with mRS grading and all-cause mortality during the 30-day period, will be summarized. Analytical methods will mirror those for primary efficacy measures.

##### 13.2.4 Safety efficacy measures:

The rate of bleeding events (severe or life-threatening, moderate, and minor bleeding) during the 30-day treatment period for the ITT population will be summarized. Safety endpoint analysis will follow the same methodology as primary efficacy measures.

##### 13.2.5 Subgroup analysis

Pre-specified subgroup analyses will explore the interaction between specific baseline characteristics and treatment effects. Baseline grouping factors for subgroup analysis are listed in Appendix 5.

#### 14 Quality Assurance and Monitoring

##### 14.1 Preservation and monitoring of original information/files

The original files are the basis for the true existence of the participants and the reliable facts of the collected data. The original files must be retained at each research center, with EDC data mirroring original files. Any discrepancies should be duly explained. The following data entered into the EDC should be derived from original documents:

- General information of the participants (initials, sex, date of birth, height, and weight).
- The date on which the participant participated in this experiment.
- Follow-up dates for the participants.
- Medical history (accompanied by disease, beginning, end, and changes).
- Medication history (treatment and medication taken, start, end, changes).
- Adverse events (occurrence, end, and change of adverse events).
- Serious adverse events (occurrence, end, and change of serious adverse events).
- Laboratory test results.
- Electrocardiography results and other medical assessments.

The investigator must allow the sponsor to conduct monitoring and audits, ethics committee reviews, and inspections by relevant regulatory agencies; the above persons should be provided access to all relevant original materials/documents.

##### 14.2 Guarantee of independence of test and control groups.

After randomization is completed, treatment teams will adhere strictly to their assigned protocols. In facilities with both test and control teams, patients will be admitted to separate wards to prevent interference. A designated hospital research supervisor will ensure protocol adherence and group independence.

##### 14.3 Pre-study work

Before the first participant is enrolled in the study, a representative of the study sponsor should visit the study center to:

1) Determine the adequacy of research center facilities.

2) Determine whether there are suitable selected patients at the study site.

3) Discuss with the researcher/center director (and other research-related personnel) their responsibilities for the research protocol and the responsibilities of the study initiator or their representative, then document these discussions in the clinical study protocol.

##### 14.4 Training of Research Center Personnel

Standardized training and evaluation for principal and sub-center researchers will be provided before enrolling the first participant. Study initiation at sub-centers is contingent upon successful evaluation. Training records will be maintained by the principal investigator.

##### 14.5 Research Monitoring

The research organizer will maintain regular contact with the research center during the research period, including:

1) Visiting research centers to provide information and support.

2) Confirm that the facilities remain compliant.

3) Confirm that all research personnel comply with the study protocol, with timely and accurate documentation of data on the EDC.

4) Conduct monthly data quality meetings, compare data on the EDC with the participant's hospital records and other records relevant to the study, including review of the trial participant's informed consent form. Direct reference to all original records for each patient (e.g., hospital records) is required.

If researchers or other staff members of the research center require information and advice concerning implementation of the study, they can contact the study initiator during the follow-up period.

##### 14.6 Archival of research documents

Researchers should adhere to the principles outlined in the clinical research protocol.

##### 14.7 Research progress

If research procedures deviate from GCP requirements or recruitment is prolonged, the study at the affected center may be terminated, and the study sponsor may prematurely end the study for safety reasons.

#### 15 Study Committee

##### 15.1 Steering Committee

Chairman: Prof Xinjian Yang, Departments of Neurosurgery and Interventional Neuroradiology, Beijing Tiantan Hospital and Beijing Neurosurgical Institute and, Capital Medical University, Beijing, China.

Members: Prof, David M Hasan, Departments of Neurosurgery, Duke

University Medical Center, Durham, NC, USA.

Prof Hongqi Zhang, Departments of Neurosurgery, Beijing XuanWu Hospital, Capital Medical University, Beijing, China.

Prof Jun Wang, Department of Neurology, Chinese PLA General Hospital, Beijing, China.

Prof Anxin Wang, China National Clinical Research Center for Neurological Diseases, Beijing Tiantan Hospital, Capital Medical University, Beijing, China.

✓ The steering committee will provide scientific and strategic direction for the trial and will have overall responsibility for study design, execution, and publication.

✓ The steering committee will also be responsible for ensuring that study execution and management are of the highest quality.

✓ It will approve the protocol and the operational guidelines of the trial prior to study commencement.

✓ The steering committee will convene regularly by teleconference or face-to-face meetings to discuss and report on the progress of the study.

**15.2 Data Safety Monitoring Board**

Chairman: Prof Chuanhui Li, Department of neurosurgery, Xuanwu Hospital Capital Medical University, Beijing, China.

Members: Prof Xue Xia, China National Clinical Research Center for Neurological Diseases, Beijing Tiantan Hospital, Capital Medical University, Beijing, China.

The Data Safety Monitoring Board (DSMB)s will meet regularly and monitor the progress of the study to ensure that the study meets the highest standards of ethics and patient safety. It is composed of Academic Members, including an independent statistician, who are not otherwise involved in the trial. A DSMB charter including membership roles and responsibilities will be approved by both the DSMB and the Steering Committee before the start of the trial. Written recommendations and their rationale will be provided to the Chairs of the Steering Committee immediately after each DSMB meeting.

##### 15.3 Imaging Assessment Committee

Chairman: Prof Binbin Sui, Department of Neuroradiology, Beijing Neurosurgical Institute, Capital Medical University, Beijing, China.

Members: Prof Jing Jing, Department of Neuroradiology, Tiantan Neuroimaging Center of Excellence, Beijing, China.

Prof Shiqing Mu, Department of Neurosurgery, Beijing Tiantan Hospital and Beijing Neurosurgical Institute, Capital Medical University, Beijing, China.

Prof Chuhan Jiang, Department of Neurosurgery, Beijing Tiantan Hospital, Capital Medical University, Beijing, China.

Prof Lian Liu, Department of Neurosurgery, Beijing Tiantan Hospital, Capital Medical University, Beijing, China.

Prof Pinyuan Zhang, Department of Neurosurgery, Hebei Medical University Third Hospital, Shijiazhuang, China.

Prof Fushun Xiao, Department of Neurosurgery, Tianjin Medical University General Hospital, Tianjin, China.

##### 15.4 Platelet function monitoring Committee

Chairman: Prof Guojun Zhang, Laboratory Diagnosis Center, Beijing Tiantan Hospital, Capital Medical University, Beijing, China.

Members: Prof Limin Zhang, Laboratory Diagnosis Center, Beijing Tiantan Hospital, Capital Medical University, Beijing, China.

##### 15.5 Adverse Event Adjudication Committee

Chairman: Prof Yuanli Zhao, Department of Neurosurgery, Peking University International Hospital, Beijing, China.

Members: Prof Yang Wang, Departments of Neurosurgery, Beijing Chaoyang Hospital, Capital Medical University, Beijing, China.

Prof Liang Li, Department of Neurosurgery, Peking University First Hospital, Beijing, China.

##### 15.6 Clinical Event Committee

Chairman: Prof Hang Shu, Department of Neurosurgery, Guangdong Provincial People's Hospital, Southern Medical University, Guangzhou, China.

Members: Prof Li Li, Stroke Center, Henan Provincial People's Hospital, Zhengzhou University, Zhengzhou, China.

Prof Ying Zhang, Department of Interventional Neuroradiology, Beijing Neurosurgical Institute, Capital Medical University, Beijing, China.

#### 16 Ethics

This trial will be conducted in full compliance with the Helsinki Declaration. The research plan will receive approval from local regulatory bodies and ethics committees.

##### 16.1 ICF

Researchers are required to adhere to the laws and regulations, as well as the principles of GCP and the Helsinki Declaration, when conducting informed consent procedures to obtain informed consent. Prior to engaging in any research-related activities, researchers must provide participants (or their parents or legal guardians, where applicable) with all relevant study information in an oral and written format that is easily understandable. Participants are required to voluntarily sign the ICF before any research activities commence. If the standard informed consent procedure is not feasible, participants must be informed using alternative methods. Researchers responsible for obtaining informed consent must also sign and date the ICF. Should any new information emerge that affects a participant's willingness to continue in the study, the researcher must inform the participant promptly and obtain a newly signed ICF.

**16.2 Ethics Committee**

Before the study begins, the research protocol, any amendments, participant information/ICF documents, and any other written materials provided to participants, along with resumes and/or qualification documents of the lead researchers, must be submitted to the ethics committee. The submission to the Ethics Committee of Beijing Tiantan Hospital of Capital Medical University should clearly state the research document code, including its version, title, and/or date. Written approval from the ethics committee is mandatory before initiating clinical research.

Throughout the course of the study, researchers are obliged to report the following to the ethics committee in compliance with local regulations: suspected unexpected SAEs, significant changes to the study protocol, non-significant adjustments as defined by local regulations, new findings or procedures that could negatively impact participant safety (including new risk/benefit analyses that may influence participant follow-up), an annual written summary of the research status, and any other documents required by the local ethics committee. Revisions must not be implemented without approval, except in cases where immediate changes are necessary to mitigate risks to participants. Researchers must meticulously document all communications with the ethics committee, retaining all records in the research files and providing copies to the sponsor.

#### Appendix 1. Definition of Ischemic Events and Vascular Events

| | |
|---|---|
| Stent thrombosis | Criteria: 1) cerebral arteriography confirms thrombosis at the stent site or distal cerebral blood vessels after stent implantation, showing an elliptical, elongated, or slightly irregular low-density image surrounded by contrast agent, and a small amount of contrast remains at the thrombus site and proximal end after contrast agent dispersal. |
| Ischemic stroke | Acute focal cerebral or retinal infarction. Criteria: (1) Clinical signs or imaging evidence of a new focal neurological defect with acute onset lasting more than 24 hours, excluding other non-ischemic causes (e.g., cerebral infection, cerebral trauma, cerebral tumor, epileptic seizure, severe metabolic disease, degenerative neurological disease, and drug side effects); or (2) Acute cerebral or retinal ischemic events, excluding other non-ischemic causes, with focal symptoms or signs lasting less than 24 hours but accompanied by radiographic evidence of new infarcts; or (3) progression of original vascular ischemic stroke (i.e., an increase in National Institutes of Health Stroke Scale score of ≥4 on the basis of primary ischemic stroke, excluding post-infarct hemorrhagic transformation or symptomatic intracranial hemorrhage) lasting longer than 24 hours with new ischemic changes on head magnetic resonance imaging or computed tomography. Etiological subtyping will be conducted in accordance with TOAST standards. |
| Transient ischemic attack | A neurological deficit of sudden onset, resolving completely, attributed to focal brain or retinal ischemia without evidence of associated acute focal infarction of the brain. Criteria: rapid onset of a focal neurological deficit that is without evidence of acute focal infarction of the brain and is not attributable to a non-ischemic etiology (brain infection, trauma, tumor, seizure, severe metabolic disease, or degenerative neurological disease). |

#### Appendix 2. GUSTO Bleeding Classification Criteria

| | |
|---|---|
| Severe or life-threatening | 1) Intracerebral hemorrhage.<br>2) Resulting in substantial hemodynamic compromise requiring treatment. |
| Moderate bleeding | Requiring blood transfusion but not resulting in hemodynamic compromise. |
| Mild bleeding | Bleeding that does not meet above criteria. |

#### Appendix 3. modified Rankin Scale (mRS)

| **Description** | **Rankin Grade** |
|---|---|
| No symptoms at all | 0 |
| No significant disability despite symptoms: able to carry out all usual duties and activities | 1 |
| Slight disability: unable to carry out all previous activities but able to look after own affairs without assistance | 2 |
| Moderate disability: requiring some help, but able to walk without assistance | 3 |
| Moderately severe disability: unable to walk without assistance, and unable to attend to own bodily needs without assistance | 4 |
| Severe disability: bedridden, incontinent, and requiring constant nursing care and attention | 5 |

#### Evaluation of mRS

The modified Rankin Scale is used to measure the results of patients' functional recovery after stroke. Bold typeface shows the formal definition of each level. The italics provide further guidance to reduce possible errors between observers, but there are no requirements for interview structure. Please note that only symptoms occurring since the stroke are considered. If the patient can walk using some assistive devices without outside help, the patient is able to walk independently.

If the two levels seem to be equally applicable to the patient, and further questions are unlikely to make an absolutely correct choice, the more severe level should be selected.

**0- No symptoms at all**

There may be mild symptoms, but no noticeable new functional limitations or new symptoms after the stroke event.

**1- No significant disability despite symptoms; capable of performing all usual duties and activities**

The patient experiences some stroke-related symptoms, whether physical or cognitive (e.g., affecting speech, reading, writing, body movement, sensation, vision, swallowing, or emotion) but is able to continue all work, social, and leisure activities as before the stroke. The distinguishing question between levels 1 and 2 could be, "Are there activities you regularly engaged in before the stroke that you are now unable to perform?". Activities considered "regular" occur more frequently than once a month.

**2- Slight disability; unable to perform all previous activities but capable of self-care without assistance**

The patient can no longer engage in some pre-stroke activities (such as driving, dancing, reading, or working) but can still manage daily self-care without assistance. The patient is able to dress, walk, eat, go to the bathroom, prepare simple foods, shop, and travel locally without assistance. The patient lives without supervision. It is envisioned that patients at this level can be home alone for a week or more without care.

**3- Moderate disability; requiring some help, but able to walk without assistance**

At this level, the patient is able to walk independently (using a walking aid) and perform basic tasks such as dressing, using the restroom, and eating. However, more complex tasks require assistance, such as shopping, cooking, or cleaning. Regular visits from someone are necessary to ensure these activities are completed. Assistance extends beyond physical care to include advice, such as managing finances, indicating a need for supervision or encouragement.

**4- Moderately severe disability; unable to walk without assistance, and unable to attend to own bodily needs without assistance**

The patient requires assistance with daily living activities, including walking, dressing, using the restroom, and eating. They need care at least once, usually twice or more daily, or must live very close to a caregiver. To differentiate levels 4 and 5, consider the patient's ability to live alone safely for any part of the day.

**5- Severe disability: bedridden, incontinent, and requiring constant nursing care and attention**

Although trained nurses are not required, the patient requires frequent care throughout the day and night.

### Appendix 4. Platelet function test guided antiplatelet therapy in the test group

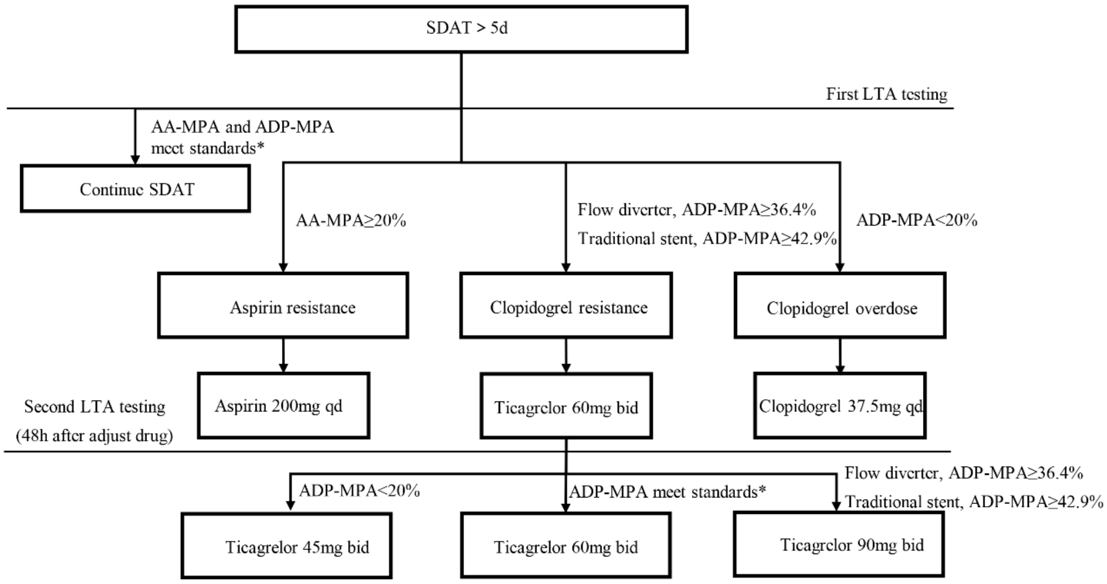

- SDAT, standard dual antiplatelet therapy (aspirin 100mg and clopidogrel 75mg daily); AA-MPA, maximum platelet aggregation rate induced by arachidonic acid, ADP-MPA, maximum platelet aggregation rate induced by adenosine diphosphatase.
- *Aspirin meets the standard: 0<AA-MPA<20%; Clopidogrel meets the standard: 20% <ADP-MPA<36.4% (for flow diverter ) or 20%<ADP-MPA<42.9% (for traditional stent).

#### Appendix 5. Description of Subgroup Types and Definitions

| Subgroups | Number of levels | Levels | |
|---|---|---|---|
| 1 | Age (Years) | 2 | > 65;<br>≤ 65 |
| 2 | Sex | 2 | Women;<br>Men |
| 3 | Body Mass Index | 2 | >30;<br>≤ 30 |
| 4 | Smoking | 2 | Yes;<br>No |
| 5 | Aneurysm size | 2 | > 7mm<br>≤ 7 mm |
| 6 | Procedure time | 2 | > 2 h<br>≤ 2 h |
| 7 | Stent type | 2 | Flow diverter;<br>Traditional stent |
| 8 | Prior ischemic stroke | 2 | Yes;<br>No |
| 9 | Atherosclerotic lesions | 2 | Yes;<br>No |

**Summary of protocol amendments**

**Protocol changes version 1.0 (May 12, 2022) to 1.1 (March 01, 2023)**

| Protocol Version 1.0 | Protocol Version 1.1 |
|---|---|
| **Study title:** Standardization of Antiplatelet Therapy in Intracranial Aneurysm Intervention with Stents: A Cluster-Randomized Controlled Cohort Study | **Study title:** Guided versus standard antiplatelet therapy in patients undergoing interventional treatment of unruptured intracranial aneurysms using stents (GATITIA): a cluster-randomized controlled cohort study |
| **Study objective:** To evaluate whether antiplatelet therapy guided by turbidimetric maximum platelet aggregation (LTA) can reduce ischemic complications in patients with unruptured intracranial aneurysms receiving stents. | **Study objective:** To determine whether platelet function test guided antiplatelet therapy reduces the incidence of ischemic complications in patients undergoing endovascular intervention for intracranial aneurysms. |
| **Primary endpoint:** All new ischemic stroke events during surgery and within 30 days after surgery. | **Primary endpoint:** Cerebral ischemic events within 30 days post-procedure |

| | |
|---|---|
| **Exploratory outcome endpoint:** | **Exploratory outcome endpoint:**<br>1) Cerebral ischemic events within 7 days post-procedure.<br>2) Modified Rankin scale score at 30 days post-procedure.<br>3) All-cause mortality within 30 days post-procedure. |
| **Safety endpoints:**<br>1) Symptomatic or asymptomatic intracranial bleeding events (including subarachnoid hemorrhage and intracerebral hemorrhage) within 30 days after surgery.<br>2) Non-intracranial bleeding events within 30 days after surgery (such as skin petechiae, ecchymoses, Nose bleeding, gum bleeding, gastrointestinal bleeding, urinary tract bleeding, etc.). | **Safety endpoints:**<br>All bleeding events within 30 days of the procedure.<br>1) severe or life-threatening bleeding—intracranial bleeding or hemodynamically impaired bleeding requiring intervention.<br>2) moderate bleeding—bleeding requiring blood transfusion but not causing hemodynamic impairment.<br>3) minor bleeding—bleeding that does not meet the criteria for severe or moderate bleeding. |

### Guided versus standard antiplatelet therapy in patients undergoing interventional treatment of unruptured intracranial aneurysms using stents

### (GATITIA)

#### Statistical Analysis Plan

**Principal Investigator: Xinjian Yang, MD, Professor of Neurointervention**

**Sponsor: Beijing Tiantan Hospital, Capital Medical University, Beijing, China**

**Version No.: V1.0**

**Date: February 01, 2023**

#### 1. Introduction

This statistical analysis plan documents the planned statistical analyses for the GATITIA study and is based on the protocol, together with any subsequent amendments. This statistical analysis plan predefines the statistical analysis population, variables, and analysis methods before database lock to ensure the reliability of the study results.

#### 2. Study design and randomization

Guided Antiplatelet Therapy in Interventional Treatment of Intracranial Aneurysms (GATITIA) is a prospective, multicenter, open-label cluster-randomized clinical trial with blinded outcome assessment. This study will enroll 590 eligible unruptured intracranial aneurysm patients with stent placement (including stent-assisted coiling and flow diversion) across 16 treatment teams in eight hospitals. The trial will last approximately 4 months at intervention centers, with treatment teams randomized 1:1 to either platelet function test-guided antiplatelet treatment (test group) or standard dual antiplatelet treatment without platelet function testing (control group).

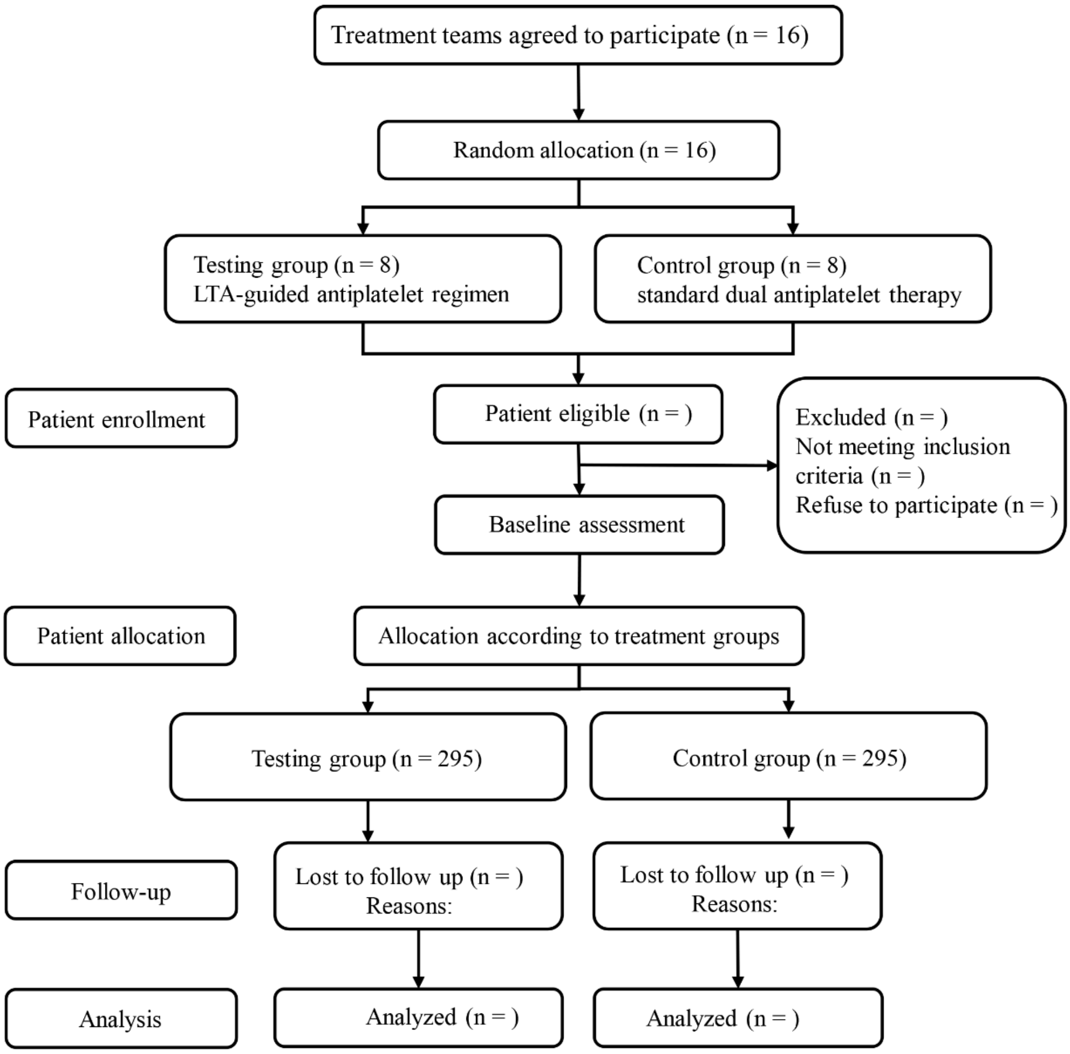

Patients in the control group will undergo stent placement without any platelet function testing, and standard dual antiplatelet preparation will be continued after the procedure. Patients in the test group will undergo platelet function testing 1 day before intracranial stent placement. Patients in the test group will receive a guided antiplatelet regimen based on light transmittance aggregometry (LTA) test results; patients with unsatisfactory platelet aggregation rates will undergo appropriate adjustments to their treatment regimen. Patients in the test group will undergo drug adjustment ≥24 hours before stent implantation. LTA test results will be re-assessed at 48 hours after any adjustment. Patients with an adequate response to antiplatelet therapy or low platelet reactivity to clopidogrel 75 mg and aspirin 100 mg will not undergo further adjustments.

#### 3. Study objective(s)

##### Primary objective

The primary objective of this trial is to evaluate whether platelet function test guided antiplatelet therapy reduces the incidence of ischemic complications in patients undergoing endovascular intervention for intracranial aneurysms, using LTA to inform the adoption of a guided antiplatelet preparation for patients with suboptimal responses to aspirin and/or clopidogrel. This study will assess the effects of LTA-guided antiplatelet treatment (double doses of aspirin or ticagrelor) during the early postoperative period (30 days), compared with standard dual antiplatelet treatment (aspirin and clopidogrel) in terms of reducing the incidence of new ischemic events in patients with unruptured intracranial aneurysms after stent placement.

##### Exploratory objectives

1) To evaluate the difference in the rate of ischemic events within 7 days between the two groups.

2) To assess the difference in modified Rankin scale (mRS) score at 30 days.

3) To evaluate the difference in all-cause mortality within 30 days between the two groups.

**Safety objective**

To evaluate the difference in the proportion of all-cause mortality within 30 days between the two groups.

The safety of the two groups will be also compared in terms of: severe or life-threatening bleeding, moderate bleeding, or minor bleeding, according to the GUSTO grading criteria.

**Other objectives**

To assess whether differential treatment effects occur in predefined subgroups:

- Age (>65 years vs ≤65 years);
- Sex (female vs male);
- Body mass index (>30 kg/m$^2$ vs ≤30 kg/m$^2$);
- Smoking (yes vs no);
- Aneurysm size (>7 mm vs ≤7 mm);
- Procedure time (>2 hours vs ≤2 hours);
- Stent type (flow diverter vs traditional stent-assisted coil embolization);
- Prior ischemic stroke (yes vs no);
- Atherosclerotic lesions (yes vs no).

#### 4. Study Efficacy Measures

##### Primary Efficacy Measure

Incidence of ischemic events, comprising a composite of stent thrombosis, ischemic stroke, and transient ischemic attack occurring within 30 days of stent placement. Stent thrombosis is defined as thrombosis at the stent site or distal cerebral vessels confirmed by digital subtraction angiography after stent implantation. Ischemic stroke is defined as rapid onset of a new focal neurological deficit with clinical or radiographic evidence of infarction, or rapid deterioration of an existing focal neurological deficit caused by a new infarction.[3] Transient ischemic attack is defined as transient neurological dysfunction caused by focal cerebral ischemia.[2]

**Exploratory Efficacy Measure**

Incidence of ischemic events within 7 days.

mRS score at 30 days post-procedure.

All-cause mortality within 30 days.

**Safety Efficacy Measure**

Rate of any bleeding event incidence (GUSTO criteria).[13]

1) Severe or life-threatening bleeding—intracranial bleeding or hemodynamically impaired bleeding requiring intervention.

2) Moderate bleeding—bleeding requiring blood transfusion but not causing hemodynamic impairment.

3) Minor bleeding—bleeding that does not meet the criteria for severe or moderate bleeding.

#### 5. Statistical Hypotheses

The primary outcome for this study is the incidence of ischemic events within 30 days. In this study, the null hypothesis of no difference in the rate of the ischemic event within 30 days between the two groups will be tested using a two-sided test with a 5% level of significance.

$H_0$: $\lambda1=\lambda2$

$H_1$: $\lambda1\neq\lambda2$

Where λ1 is the rate of the ischemic event within 30 days in the test group and λ2 is the same outcome in the control group.

#### 6. Sample size calculation

In a previous randomized controlled study of patients undergoing stent placement for intracranial aneurysm treatment, the incidences of ischemic events within 30 days of treatment in patients who received antiplatelet drug adjustment based on platelet function testing and patients who received conventional treatment without monitoring were 5.1% and 12.1%, respectively.[20] Based on a previous study, the intracluster correlation coefficient was assumed to be 0.002 in this study.[19] Using a test level of 0.05 and a power of 0.8, it is anticipated that 35 patients will be recruited from each cluster (treatment team), requiring a total of 560 patients. Considering a 5% attrition rate, the recruitment target is set at 590 patients (295 per group). Each group will randomly have one treatment team that recruits 36 patients; the other teams will recruit 37. PASS14 (NCSS, LLC) will be used to calculate the sample size.

#### 7. Analysis Populations

SPSS software, version 26.0 (IBM Corp., Armonk, NY, USA) and Prism software (GraphPad Prism 9.0 software) will be utilized to perform statistical analyses of efficacy and safety measures. The level of statistical significance will be set at $P<.05$.

According to the basic principles of intention-to-treat analysis (ITT), all participants who undergo randomization and stent placement will be included in the full analysis set.

#### 8 Protocol Deviations

Participant data will be reviewed for evidence of protocol violations to ensure adherence to the study protocol. Detailed inclusion and exclusion criteria are outlined in the protocol. Participants who deviate from the protocol by changing treatments without authorization or engaging in prohibited concurrent treatments will be excluded from the per-protocol analysis. A preliminary list of potential protocol violators will be compiled for clinical assessment. The research team will finalize this list to determine which participants should be excluded from the per-protocol population.

#### 9. Demography and baseline characteristics

The following demographic information will be listed and summarized for participants in each treatment group: age, sex, body mass index, hypertension, diabetes, coronary heart disease, hyperlipidemia, smoking and drinking history, complete blood count, liver function, kidney function, blood glucose, blood lipids, coagulation parameters, LTA, modified Rankin scale (mRS), aneurysm characteristics, aneurysm characteristics, and diameter of parent artery.

Continuous data will undergo normality testing with the Shapiro–Wilk test. Normally distributed data will be presented as means ± standard deviations, and skewed data as medians with interquartile ranges. Categorical data will be reported as counts and percentages. Baseline differences between groups will be analyzed using the independent Student's t-test or Mann–Whitney U test for continuous variables, and the Pearson $\chi^2$ test for categorical variables.

#### 10. Analysis of efficacy outcomes

##### Primary efficacy analysis

The primary endpoint is the rate of ischemic events within the 30-day treatment period. The efficacy analysis will include all randomized participants who completed stent placement.

##### Main Analyses

The incidence of ischemic events (stent thrombosis, ischemic stroke, transient ischemic attack) during the 30-day period for the intention-to-treat (ITT) population will be summarized. Analysis will employ generalized linear mixed-effects models with intervention as the fixed effect and treatment team as the random effect to address potential intra-team outcome correlations. Unbalanced variables between groups (P<.1) will be incorporated into the model to adjust for confounders. The frequency of each ischemic event will be calculated for each group. Odds ratios with 95% confidence intervals (CIs) will be reported.

**Interactions with subgroups**

Summary tables will be produced for the predefined subgroups. Interactions between the two main groups and these subgroups will be investigated using generalized linear mixed-effects models, with intervention as the fixed effect and treatment team as the random effect, to adjust for potential correlations in outcomes within each treatment team. A separate model will be used for each interaction to determine its statistical significance. These data will also be presented graphically on a tree plot.

The predefined subgroups including:

➢ Age (>65 years vs ≤65 years);

➢ Sex (female vs male);

➢ Body mass index (>30 kg/$m^2$ vs ≤30 kg/$m^2$);

➢ Smoking (yes vs no);

➢ Aneurysm size (>7 mm vs ≤7 mm);

- Procedure time (>2 hours vs ≤2 hours);
- Stent type (flow diverter vs traditional stent-assisted coil embolization);
- Prior ischemic stroke (yes vs no);
- Atherosclerotic lesions (yes vs no).

**Exploratory efficacy analyses**

**Incidence of ischemic events within 7 days**

The ischemic event rate within the 7 days will be analyzed using a generalized linear mixed-effects model, with intervention as the fixed effect and treatment team as the random effect. Odds ratios with 95% CIs will be reported.

**mRS score at 30 days**

Differences between test and control group in the mRS score within the 30 days will be analyzed using a generalized linear mixed-effects models, with intervention as the fixed effect and the treatment team as the random effect. Odds ratios with 95% CIs will be reported.

**All-cause mortality within 30 days**

The mortality rate within the 30 days will be analyzed using a generalized linear mixed-effects models, with intervention as the fixed effect and the treatment team as the random effect. Odds ratios with 95% CIs will be reported.

#### 11. Safety analyses

**Adverse events**

AEs will be grouped according to system organ class. Separate data display listings and summaries will be presented for AEs that begin prior to the first dose of study medication (pretreatment), during administration of study medication (during treatment), and after the last dose of study medication (post-treatment).

Within each group, the number and percentage of patients experiencing an AE will be summarized by system organ class and preferred term; Fisher's exact test will be used to compare the numbers of grouped AE events between treatment groups. A separate summary will also be provided for AEs experienced by more than 5% of patients in either group.

**Deaths and serious adverse events**

Summary tables and data displays will be provided for serious adverse events. All deaths and serious AEs will be documented in a case narrative format in the clinical study report. The number of deaths that occur during the treatment period will be summarized; Fisher's exact test will be used to compare the numbers of deaths between groups.
